# Using Twitter Data Analysis to Understand the Perceptions, Awareness, and Barriers to the Wide Use of Pre-Exposure Prophylaxis in the United States

**DOI:** 10.1101/2022.12.19.22283677

**Authors:** Arslan Erdengasileng, Shubo Tian, Sara S. Green, Sylvie Naar, Zhe He

**Affiliations:** Department of Statistics, Florida State University Tallahassee, USA; Department of Behavioral Sciences and Social Medicine, Florida State University, Tallahassee, USA; School of Information, Florida State University Tallahassee, USA

**Keywords:** Social Media, Natural Language Processing, HIV

## Abstract

User-generated social media posts such as tweets can provide insights about the public’s perception, cognitive, and behavioral responses to health-related issues. Pre-Exposure Prophylaxis (PrEP) is one of the most effective ways to reduce the risk of HIV infection. However, its utilization is low in the US, especially among populations disproportionately affected by HIV such as the age group of under 24 years old. It is therefore important to understand the barriers to the wider use of PrEP in the US using social media posts. In this study, we collected tweets from Twitter about PrEP in the past 4 years to identify such barriers by first identifying tweets about personal discussions, and then performing textual analysis using word analysis, UMLS semantic type analysis, and topic modeling. We found that the public often discussed advocacy, risks/benefits, access, pricing, insurance coverage, legislation, stigma, health education, and prevention of HIV. This result is consistent with the literature and can help identify strategies for promoting the use of PrEP, especially among young adults.

## 1. Introduction

Human immunodeficiency virus (HIV) continues to be a public health concern in the United States, with increased risk for minority and other vulnerable populations [1]–[4]. Young people in particular, especially those who identify as sexual and gender minority (SGM) and/or are of a racial minority, account for a large proportion of newly diagnosed HIV cases[3], indicating a significant need for tailored and targeted measures aimed at prevention within this population.

One of the primary prevention strategies, approved by the US Food and Drug Administration since 2012, is pre-exposure prophylaxis (PrEP); a once daily tablet regimen available through prescription, that has been shown to be highly effective at decreasing the rate of HIV transmission, particularly when made available to high-risk populations [2]–[4]. However, large-scale studies have uncovered a concerningly low level of PrEP awareness and usage, particularly among these groups [1], [5].

In attempt to address this, institutional and community-based initiatives, provider education efforts, and multimedia campaigns have attempted to spread awareness of the risks of HIV, the importance of prevention, and methods for prevention such as PrEP, but in some instances these efforts have resulted in issues related to misinformation and stigma, which have negatively impacted perceptions of PrEP as a safe and effective measure of prevention [5], [6]. Additionally, concerns about access and low-quality patient/provider relationships have also been attributed to reduced uptake [5], [6].

Social media has been widely used for HIV research for promoting HIV prevention [3], [7], recruiting participants for research studies [8], surveillance [9], [10], and secondary data analysis [11]. For example, Heerden et al. [9] used the prevalence and patterns of social media use related to HIV risk in South Africa using Twitter, Instagram, and YouTube. They also mapped and statistically tested the differences in HIV-related posts in different regions of South Africa. Young et al. [10] collected over 550 million tweets related to sexual behaviors and drug use and merged the data with AIDSVU on HIV cases, and then performed negative binomial regression analysis to assess the relationship between HIV risk tweeting and prevalence of HIV by county, while controlling for socioeconomic status measures. Liu and Lu [11] analyzed the psychology, behavior, and demand of online HIV population and examined the network community structure using the data from “HIV bar”, the largest bar related to HIV on Baidu Tieba platform. They reported that the closer the social distance between members of the community, the more similar their topics. They also found that those who suspected to have HIV infection or first in contact with high-risk behaviors tend to ask for help and advice on social networking sites, rather than immediately going to a hospital for blood tests.

In line with similar efforts to understand awareness surrounding PrEP and its prescription brand name drug Truvada, we attempt to gather insight through the analysis of discussion on one of the most popular social media platform - Twitter. Twitter, ubiquitously used by adolescents and young adults as a means of communication and information gathering and sharing, may provide some insight into the current levels of awareness, perceptions, and attitudes surrounding PrEP usage, as well as uncover areas wherein researchers, providers, and public health officials may address gaps in awareness and education. In this study, we use various natural language processing and text mining techniques to uncover these areas at different levels (i.e., words, semantics, topic). This study aims to answer the following research questions: RQ1: what are the most popular topics in personal discussion tweets? RQ2: how the frequency of words and semantic types differ between promotional tweets and personal discussion tweets? RQ3: how the discussion tweets and promotional tweets vary over time?

## II. Method

## A. Overview of the Method

In this study, we first retrieved tweets related to PrEP and Truvada from Twitter. Because the collected data contain a mix of tweets about promotional information of PrEP/Truvada, personal discussion of PrEP, and tweets that are not about PrEP/Truvada at all, it is important to distinguish between these tweets for subsequent analysis. We employed deep-learning-based text classification to categorize tweets into three such categories: personal discussion, promotional information, or irrelevant tweets. To train and test a text classification model, we randomly selected a set of tweets and two Pre-Med undergraduate students manually annotated them. We then built and evaluated deep learning-based text classifiers and selected the best one to classify all the other unannotated tweets after pre-processing. We then performed word analysis and semantic type analysis on both personal discussion and promotional tweets. To uncover the major topics of personal discussion, we performed topic modeling on them. The detailed process is shown in Figure 1.

**Fig. 1.**
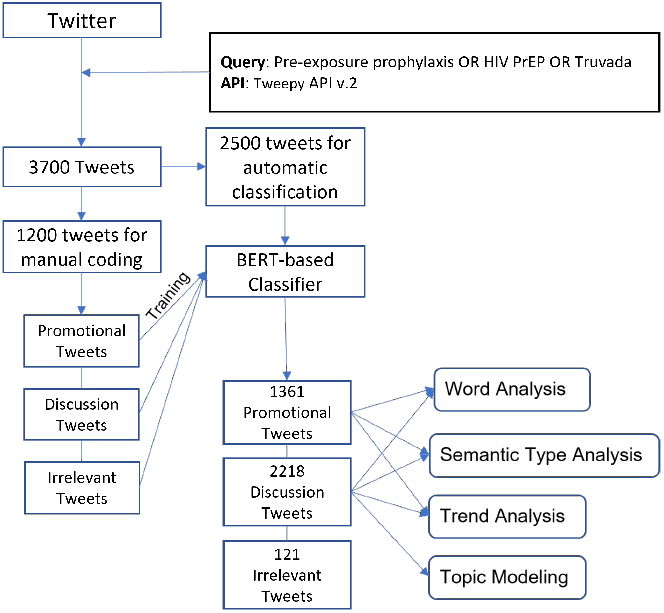
Flow chart of overall method

### B. Data Collection, Annotation and Pre-processing

Tweets related to PrEP and Truvada were curated from Twitter using the Tweepy API v2. We queried tweets that contained any of the three keywords of ‘Pre-exposure prophylaxis’, ‘HIV PrEP’ or ‘Truvada’. We searched for tweets that were posted from 1/1/2018 to 1/1/2022, and limited our search on tweets in English and in the US. Retweets were also excluded. In the end, we curated 3,700 tweets for our study.

After relevant tweets were retrieved, 1,200 tweets were randomly selected for manual annotation. Two Pre-Med annotators SH and JV annotated the 1,200 tweets into 3 categories. First off, an annotation guideline was developed by ZH and AE and 200 tweets were selected for independent annotation by each annotator following the guideline. SH and JV reached an inter-rater agreement of a Cohen Kappa score of 0.7056, indicating substantial agreement [12]. To improve the agreement, both annotators reviewed 100 tweets together and then re-annotated these 200 tweets. In the end, two annotators reached an inter-rater agreement of a Cohen Kappa score of 0.8029, indicating almost perfect agreement. Then they separately annotated 500 tweets each. In total, 1,200 tweets were manually categorized into three aforementioned categories. These 12,00 tweets were used as the labelled data to train and test the text classification models.

Before tweet classification, word and semantic type analysis, and topic modeling, the retrieved tweets were first pre-processed in the following six steps: (1) removing all the emojis; (2) replacing the usernames by “USER”; (3) replacing the URL links by “URL”; (4) replacing ‘&amp’ by an empty string; (5) removing all the ‘#’ signs; (6) replacing multiple spaces and newlines with a single space.

### C. Tweet Classification

In this study, we are interested in the tweets relevant to Truvada/PrEP. However tweets retrieved by keyword queries can include irrelevant ones. To extract the relevant tweets, we categorized the collected tweets into three categories of irrelevant, promotional and personal discussion tweets. Irrelevant tweets are those with content completely irrelevant to Truvada/PrEP (e.g., “prep for an exam”). Promotional tweets are those containing promotional information about Truvada/PrEP, such as advertising, sales promotion, and public relations.

Personal discussion tweets are those talking about personal health experiences, concerns with Truvada/PrEP. To group the retrieved tweets into different categories, we first manually annotated a subset of the tweets and then built multiple BERT-based classifiers to classify the remaining tweets. We compared the performance of different classifiers in terms of a balance between micro F1 and F1, and selected the one with best performance for classification of the remaining tweets.

Bidirectional Encoder Representations from Transformers (BERT) model [13] has been recognized as the state-of-the-art model for text classification. BERT uses Transformer, an attention mechanism that discovers contextual relationships between words (or subwords) in the text. Transformer is the component of the model that gives BERT its improved ability to comprehend linguistic ambiguity and context.

BERT models further pre-trained with domain specific corpus usually perform even better on domain specific datasets. Since our curated tweets were focused on health information about HIV, Truvada and PrEP, we built several BERT-based classifiers using different BERT models further pre-trained with biomedical corpus and compared their performance to the classifier based on the BERT-base model.

We chose BERT-base model as our baseline model and selected several other BERT models including BERT-large [13], BioBERT [14], BlueBERT [15], PubMedBERT-abstract-only [16], and PubMedBERT-fulltext [16] for tweet classification in this study.

The BERT-base model was pre-trained on BooksCorpus and the English Wikipedia and has 12 layers with 12 bidirectional self-attention heads in each layer.

The BERT-large model was also pre-trained on BooksCorpus and English Wikipedia. Only difference is that it has 24 layers with 16 bidirectional self-attention heads in each layer. The BioBERT model [14] was initialized with weights from BERT and then further pre-trained on biomedical domain corpora (PubMed abstracts and PMC full-text articles).

The BlueBERT model was initialized with weights from BERT and pre-trained using PubMed abstracts and MIMIC-III notes [15].

The PubMedBERT model was pre-trained on biomedical text from scratch (PubMed abstract only or PubMed full-text). This is different from BioBERT and BlueBERT which were initialized with BERT-base weights.

### D. Word Analysis and UMLS Semantic Type Analysis

After promotional and personal discussion tweets were extracted, we are interested in what words were commonly used and what were the common semantic types in each group. As such, we implemented word analysis and semantic type analysis using QuickUMLS [17]. QuickUMLS is a tool for fast, unsupervised biomedical concept extraction from medical text. It parses a text string into several spans and decides if each span has matched concepts in the UMLS by similarity algorithms such as Jaccard similarity. The matched spans can then be mapped to concept unique identifiers (CUIs) and semantic types which are associated with the correspondent matched concepts. We ran QuickUMLS on the pre-processed tweets with a Jaccard similarity score of 0.9.

There are more than 100 source vocabularies in the Unified Medical Language System (UMLS) Metathesaurus, including popular vocabularies such as MeSH, ICD-9, ICD-10, RXNORM, CPT, etc. Because the UMLS largely relied on the automated integration technique for terminology mapping, it has noises. To ensure the high quality of concept extraction, it is common to choose a small subset of the UMLS. Based on the fact that MeSH is one of the highest quality and widely used thesauri for biomedical NLP in the UMLS, we decided to use only the MeSH vocabulary for our word and semantic analysis in this study.

### E. Topic modeling

Uncovering the major topics discussed in tweets related to Truvada/PrEP can be very useful. In this study, we are interested in how the topics vary in personal discussion about Truvada/PrEP. Therefore, we applied topic modeling method to tweets in the category of personal discussion.

Topic modeling aims to assign each tweet a set of abstract topics. It is a method that groups words and phrases that best describe a group of documents automatically. To be more specific, we implemented Latent Dirichlet Allocation (LDA) statistic model [18], which is one of the most popular topic modeling algorithms and can be easily implemented with a Python package called Gensim [19], to discover the topics discussed in each tweet.

In this step, with the pre-processed tweets classified as personal discussion, we further processed the tweets in the following steps before implementing LDA topic modeling: (1) Tokenize each tweet into words and keep only the nouns; (2) Lemmatize each word to group different forms of the same word to one form; (3) Collect frequent common words as stop words and remove them; (4) Keep only words appearing more than twice in all personal discussion tweets to filter out less frequent words.

One of the major challenges in topic modeling is to decide the number of topics. A straightforward way to choose the topic number is to build multiple LDA models with different numbers of topics and pick the one with highest coherence value. However, sometimes the model with the highest coherence value is not easy to interpret. Therefore, in practice we need to select a few candidate number of topics and manually check the interpretability of the models. After manually summarizing the main topics, we might find some similar or duplicated topics that can be merged to help the topic modeling be more concise and reasonable. In this study, we performed topic modeling following the aforementioned process.

## III. Results

### A. Tweets Classification

We compared performance of different classifiers for tweet classification by experiments based on the 1,200 annotated tweets. For the experiments, we split the annotated tweets into datasets for training, validation and test at the ratio of 80:10:10, i.e., 960 tweets for training, 120 tweets for validation and 120 tweets for test. Different classifiers were built on top of different pre-trained BERT models including BERT-base, BERT-large, PubMedBERT-abstract-only, PubMedBERT-fulltext, BioBERT, and BlueBERT. We trained, validated, and tested each model with the same datasets, and used micro F1 and F1 on categories of promotional and personal discussion tweets for comparison of model performance and selection of classifier for classification of the remaining 2,500 tweets. The performance metrics of tweet classification by different classifiers are listed in Table I.

**TABLE I.** BERT-BASED CLASSIFIERS PERFORMANCE

| Model | Micro F1 | F1, Personal Discussion | F1, Promotional | F1, Irrelevant |
| --- | --- | --- | --- | --- |
| BERT-base | 0.792 | 0.85 | 0.72 | 0.55 |
| BERT-large | 0.800 | 0.85 | 0.75 | 0.4 |
| PubmedBERT-abstract-only | 0.825 | 0.89 | 0.79 | 0 |
| PubmedBERT-fulltext | 0.775 | 0.84 | 0.72 | 0.2 |
| BioBERT | 0.775 | 0.84 | 0.71 | 0 |
| BlueBERT | 0.825 | 0.88 | 0.78 | 0.25 |

As shown in Table I, our baseline model, the BERT-base model, achieved a Micro F1 score of 0.792, and 0.85 and 0.72 of F1 on categories of promotional and personal discussion tweets. The classifier based on the pre-trained PubMedBERT-abstract-only model yielded the best performance (micro F1 score of 0.825, F1 score of 0.89 for personal discussion tweets, and F1 score of 0.79 for promotional tweets). In the end, we selected the PubMedBERT-abstract-only based classifier to classify the remaining 2,500 tweets into three categories due to its superior overall performance. As such, all the 3,700 tweets were grouped into 1,361 promotional tweets (302 annotated, 1,059 classified), 2,218 personal discussion tweets (793 annotated, 1,425 classified), and 121 irrelevant tweets (104 annotated, 17 classified).

### B. Tweet Counts Over Time

Our curated 3,700 tweets were posted from January 2018 to December 2021. We compared the numbers of promotional tweets and personal discussion tweets counted every quarter. As shown in Figure 2, we had the following observations: (1) There were more personal discussion tweets over promotional tweets in nearly all the quarters (except the first one); (2) Personal discussion tweets had an increasing trend from the first quarter of 2018 to the last quarter of 2019 and an decreasing trend from the last quarter of 2019 to the last quarter of 2021 (Note that in October 2019, the FDA approved Descovy for HIV Pre-Exposure Prophylaxis [PrEP]); (3) The number of promotional tweets increases from 2018 first quarter to 2019 second quarter and decreases since then; (4) The biggest drop on the number of tweets between two categories occurred from the third quarter of 2019 to the third quarter of 2020.

**Fig. 2.**
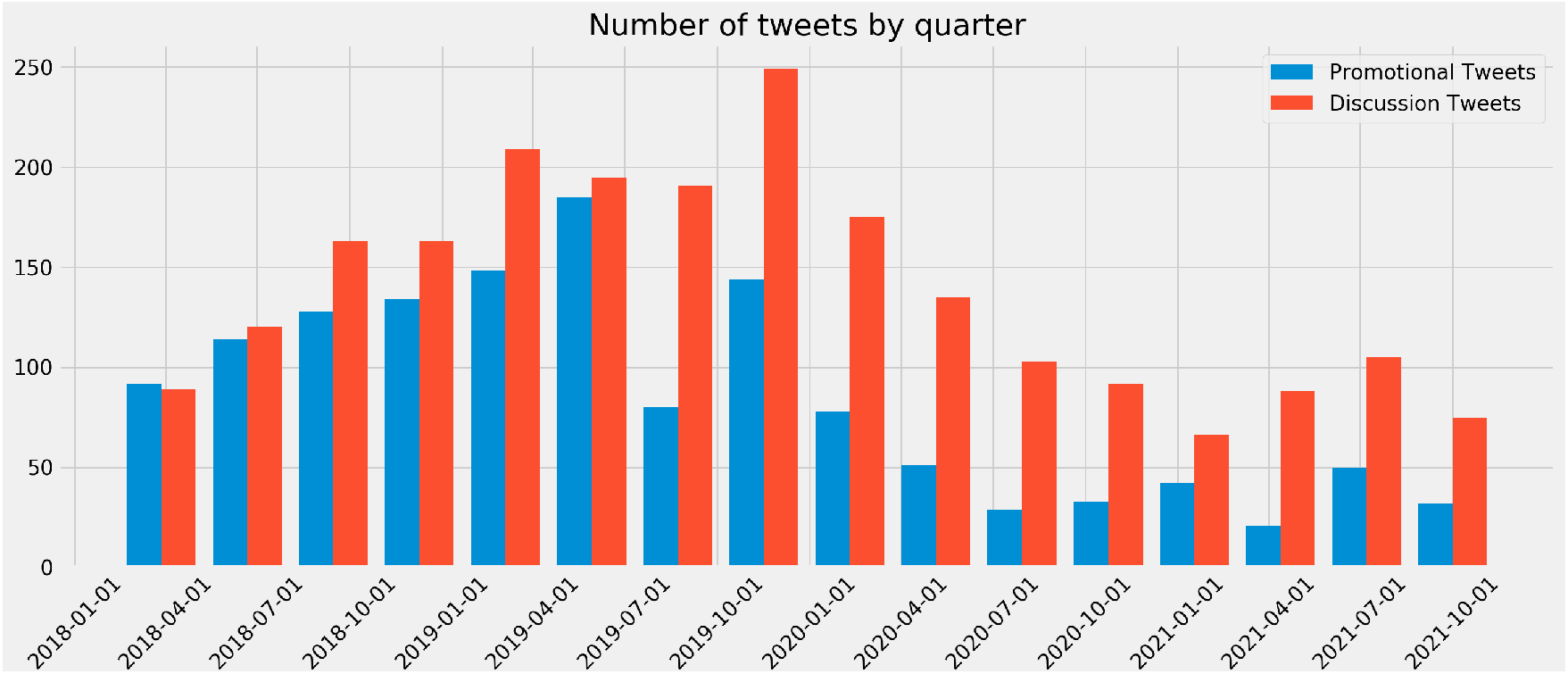
Trend analysis

### C. Word Analysis

For word analysis, we identified the top 30 words that were used in both promotional and personal discussion tweets, as shown in Table II. The frequency of a word means the number of tweets include the specific word and the percentage of a word means the frequency divided by the total number of tweets in each category. We used QuickUMLS [17] to extract biomedical concepts contained in each tweet in terms of word and kept only the words that had matched concepts in the MeSH vocabulary. In order to determine the frequency and percentage of the most frequently occurring words, we counted the number of tweets that contained a specific word. The objective of word analysis is to compare and identify word differences between the two groups of tweets.

**TABLE II.**
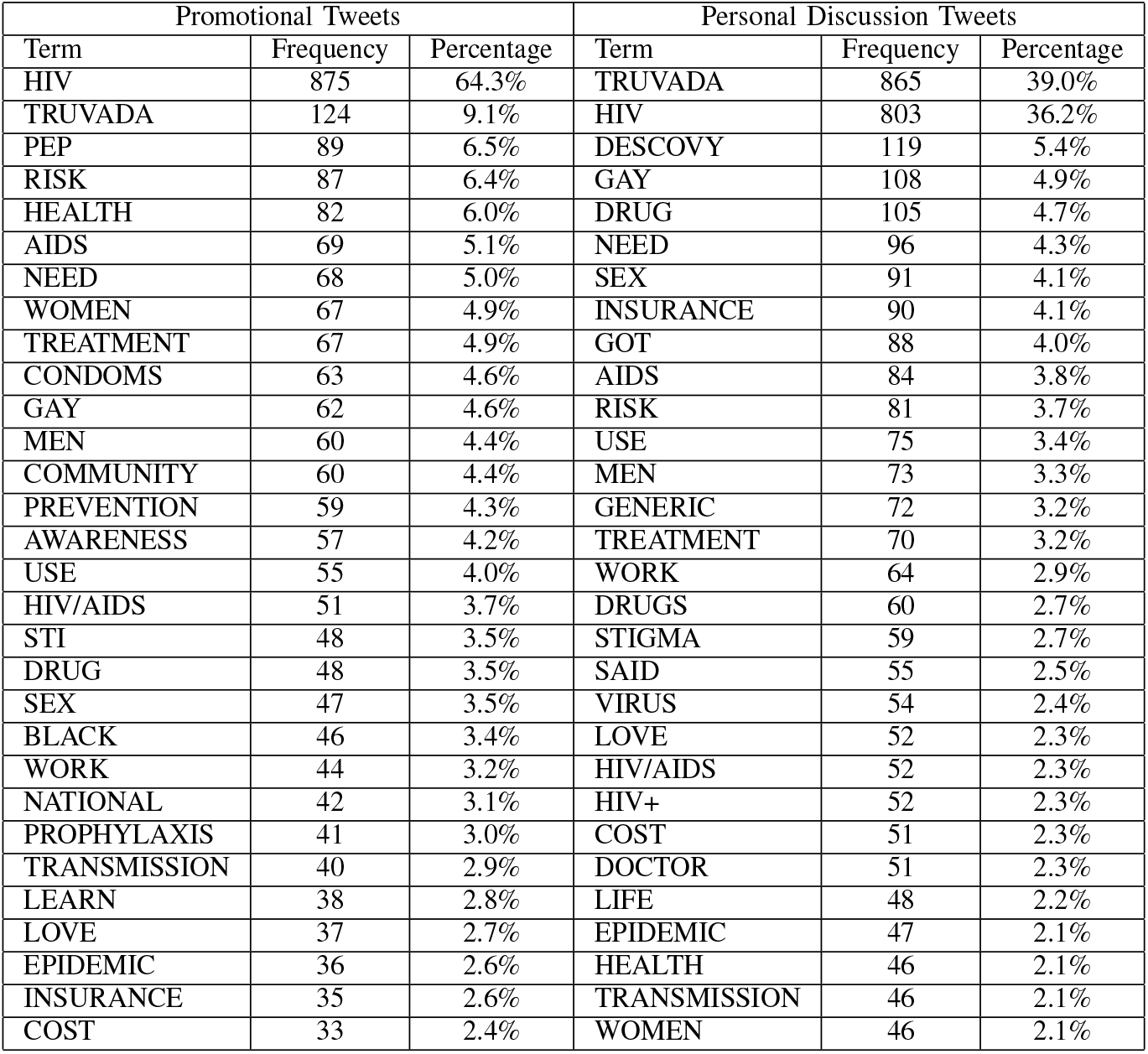
Word analysis for Promotion and Discussion

| Promotional Tweets |  |  | Personal Discussion Tweets |  |  |
| --- | --- | --- | --- | --- | --- |
| Term | Frequency | Percentage | Term | Frequency | Percentage |
| HIV | 875 | 64.3% | TRUVADA | 865 | 39.0% |
| TRUVADA | 124 | 9.1% | HIV | 803 | 36.2% |
| PEP | 89 | 6.5% | DESCOVY | 119 | 5.4% |
| RISK | 87 | 6.4% | GAY | 108 | 4.9% |
| HEALTH | 82 | 6.0% | DRUG | 105 | 4.7% |
| AIDS | 69 | 5.1% | NEED | 96 | 4.3% |
| NEED | 68 | 5.0% | SEX | 91 | 4.1% |
| WOMEN | 67 | 4.9% | INSURANCE | 90 | 4.1% |
| TREATMENT | 67 | 4.9% | GOT | 88 | 4.0% |
| CONDOMS | 63 | 4.6% | AIDS | 84 | 3.8% |
| GAY | 62 | 4.6% | RISK | 81 | 3.7% |
| MEN | 60 | 4.4% | USE | 75 | 3.4% |
| COMMUNITY | 60 | 4.4% | MEN | 73 | 3.3% |
| PREVENTION | 59 | 4.3% | GENERIC | 72 | 3.2% |
| AWARENESS | 57 | 4.2% | TREATMENT | 70 | 3.2% |
| USE | 55 | 4.0% | WORK | 64 | 2.9% |
| HIV/AIDS | 51 | 3.7% | DRUGS | 60 | 2.7% |
| STI | 48 | 3.5% | STIGMA | 59 | 2.7% |
| DRUG | 48 | 3.5% | SAID | 55 | 2.5% |
| SEX | 47 | 3.5% | VIRUS | 54 | 2.4% |
| BLACK | 46 | 3.4% | LOVE | 52 | 2.3% |
| WORK | 44 | 3.2% | HIV/AIDS | 52 | 2.3% |
| NATIONAL | 42 | 3.1% | HIV+ | 52 | 2.3% |
| PROPHYLAXIS | 41 | 3.0% | COST | 51 | 2.3% |
| TRANSMISSION | 40 | 2.9% | DOCTOR | 51 | 2.3% |
| LEARN | 38 | 2.8% | LIFE | 48 | 2.2% |
| LOVE | 37 | 2.7% | EPIDEMIC | 47 | 2.1% |
| EPIDEMIC | 36 | 2.6% | HEALTH | 46 | 2.1% |
| INSURANCE | 35 | 2.6% | TRANSMISSION | 46 | 2.1% |
| COST | 33 | 2.4% | WOMEN | 46 | 2.1% |

The most frequent word in promotional tweets is HIV (64.3%), while the most frequent words in personal discussion tweets are Truvada (39%) and HIV (36.2%). This makes sense because the query includes HIV and Truvada. However, the proportion of Truvada in personal discussion tweets is 27.1% higher than Truvada in promotional tweets.

The other frequent words in promotional tweets (the percentages are from 2.4% - 6.5%) are mainly about risks, prevention, and awareness. While in the personal discussion tweets, the rest of the words (the percentages are from 2% - 5.4%) are about drugs, treatments, insurance coverage, sex behavior, and stigma, etc.

### D. UMLS Semantic Type Analysis

Semantic type is a higher level concept that compiles similar concepts into a single category. There are 127 semantic types in UMLS. In this study, we are interested in the most prevalent semantic types. With the biomedical concepts extracted during the word analysis process, we were able to find the associated semantic types for each tweet. We can then uncover the most prevalent semantic types by counting the number of tweets associated with each semantic type. Table III lists the top 15 frequent semantic types. The ranked lists of prevalent semantic types for promotional tweets and personal discussion tweets are quite similar. Pharmacologic Substance and Disease or Syndrome are the top 2 semantic types in both groups. The percentage of promotional tweets containing terms of semantic types Disease or Syndrome, Immunologic Factor, Virus are much higher than the personal discussion tweets. This is understandable as promotional tweets may often mention such information to increase the public’s awareness.

**TABLE III.** Semantic type analysis for promotion and discussion

| Promotional Tweets |  |  | Personal Discussion Tweets |  |  |
| --- | --- | --- | --- | --- | --- |
| Semantic Type | Frequency | Percentage | Semantic Type | Frequency | Percentage |
| Pharmacologic Substance | 1044 | 76.7% | Pharmacologic Substance | 1692 | 76.3% |
| Disease or Syndrome | 1006 | 73.9% | Disease or Syndrome | 1052 | 47.4% |
| Immunologic Factor | 886 | 65.1% | Nucleic Acid, Nucleoside, or Nucleotide | 873 | 39.4% |
| Virus | 885 | 65.0% | Immunologic Factor | 835 | 37.6% |
| Population Group | 364 | 26.7% | Virus | 832 | 37.5% |
| Idea or Concept | 309 | 22.7% | Population Group | 589 | 26.6% |
| Mental Process | 258 | 19.0% | Mental Process | 417 | 18.8% |
| Intellectual Product | 211 | 15.5% | Idea or Concept | 373 | 16.8% |
| Qualitative Concept | 208 | 15.3% | Functional Concept | 309 | 13.9% |
| Functional Concept | 179 | 13.2% | Organic Chemical | 271 | 12.2% |
| Therapeutic or Preventive Procedure | 168 | 12.3% | Intellectual Product | 263 | 11.9% |
| Organic Chemical | 166 | 12.2% | Organism Function | 235 | 10.6% |
| Geographic Area | 166 | 12.2% | Qualitative Concept | 230 | 10.4% |
| Amino Acid, Peptide, or Protein | 142 | 10.4% | Amino Acid, Peptide, or Protein | 214 | 9.6% |

### E. Topic modeling

For topic modeling of tweets of personal discussion about Truvada/PrEP, first we calculated coherence values for 11 different LDA models with number of topics ranging from 5 to 15 and manually evaluated the results of different models. We found out that the most adequate number of topics for LDA topic modeling was 15. After the 15 topics were identified by the LDA model, keywords in each topic were listed and SG, an HIV researcher, determined the themes for each topic based on some sample tweets together with the keywords in the topic. Table IV shows the keywords and themes for each of the original 15 topics identified by the LDA model. Column keywords shows the most frequent words in each topic, and the column “Summary topic” was the themes summarized by ZH, AE, and SG. From this column, we can see there exist similar or duplicated topics that need to be merged.

**TABLE IV.**
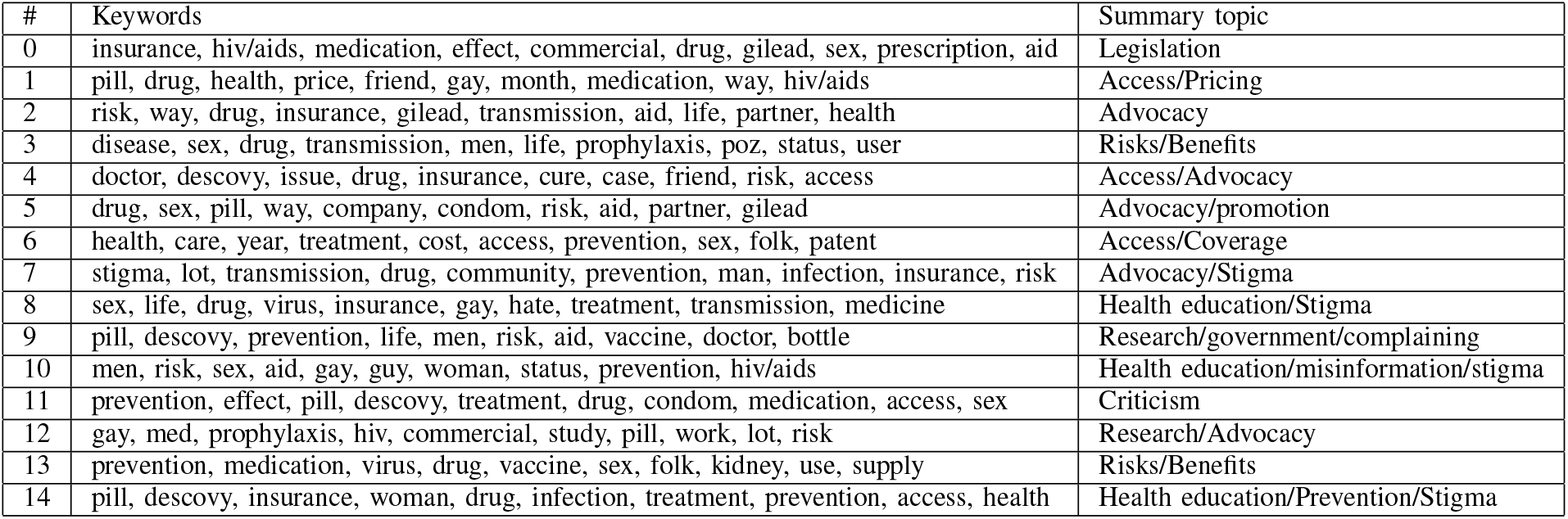
Topic Modeling

| # | Keywords | Summary topic |
| --- | --- | --- |
| 0 | insurance, hiv/aids, medication, effect, commercial, drug, gilead, sex, prescription, aid | Legislation |
| 1 | pill, drug, health, price, friend, gay, month, medication, way, hiv/aids | Access/Pricing |
| 2 | risk, way, drug, insurance, gilead, transmission, aid, life, partner, health | Advocacy |
| 3 | disease, sex, drug, transmission, men, life, prophylaxis, poz, status, user | Risks/Benefits |
| 4 | doctor, descovy, issue, drug, insurance, cure, case, friend, risk, access | Access/Advocacy |
| 5 | drug, sex, pill, way, company, condom, risk, aid, partner, gilead | Advocacy/promotion |
| 6 | health, care, year, treatment, cost, access, prevention, sex, folk, patent | Access/Coverage |
| 7 | stigma, lot, transmission, drug, community, prevention, man, infection, insurance, risk | Advocacy/Stigma |
| 8 | sex, life, drug, virus, insurance, gay, hate, treatment, transmission, medicine | Health education/Stigma |
| 9 | pill, descovy, prevention, life, men, risk, aid, vaccine, doctor, bottle | Research/government/complaining |
| 10 | men, risk, sex, aid, gay, guy, woman, status, prevention, hiv/aids | Health education/misinformation/stigma |
| 11 | prevention, effect, pill, descovy, treatment, drug, condom, medication, access, sex | Criticism |
| 12 | gay, med, prophylaxis, hiv, commercial, study, pill, work, lot, risk | Research/Advocacy |
| 13 | prevention, medication, virus, drug, vaccine, sex, folk, kidney, use, supply | Risks/Benefits |
| 14 | pill, descovy, insurance, woman, drug, infection, treatment, prevention, access, health | Health education/Prevention/Stigma |

We combined the similar or duplicated topics and ended up with 5 main topics. All the original tweets were grouped into the 5 topics and a word cloud was plotted for each group as shown in Figure 3 (excluding HIV, Truvada, PrEP). We listed brief description and typical example tweets for each topic as follows.

**Fig. 3.**
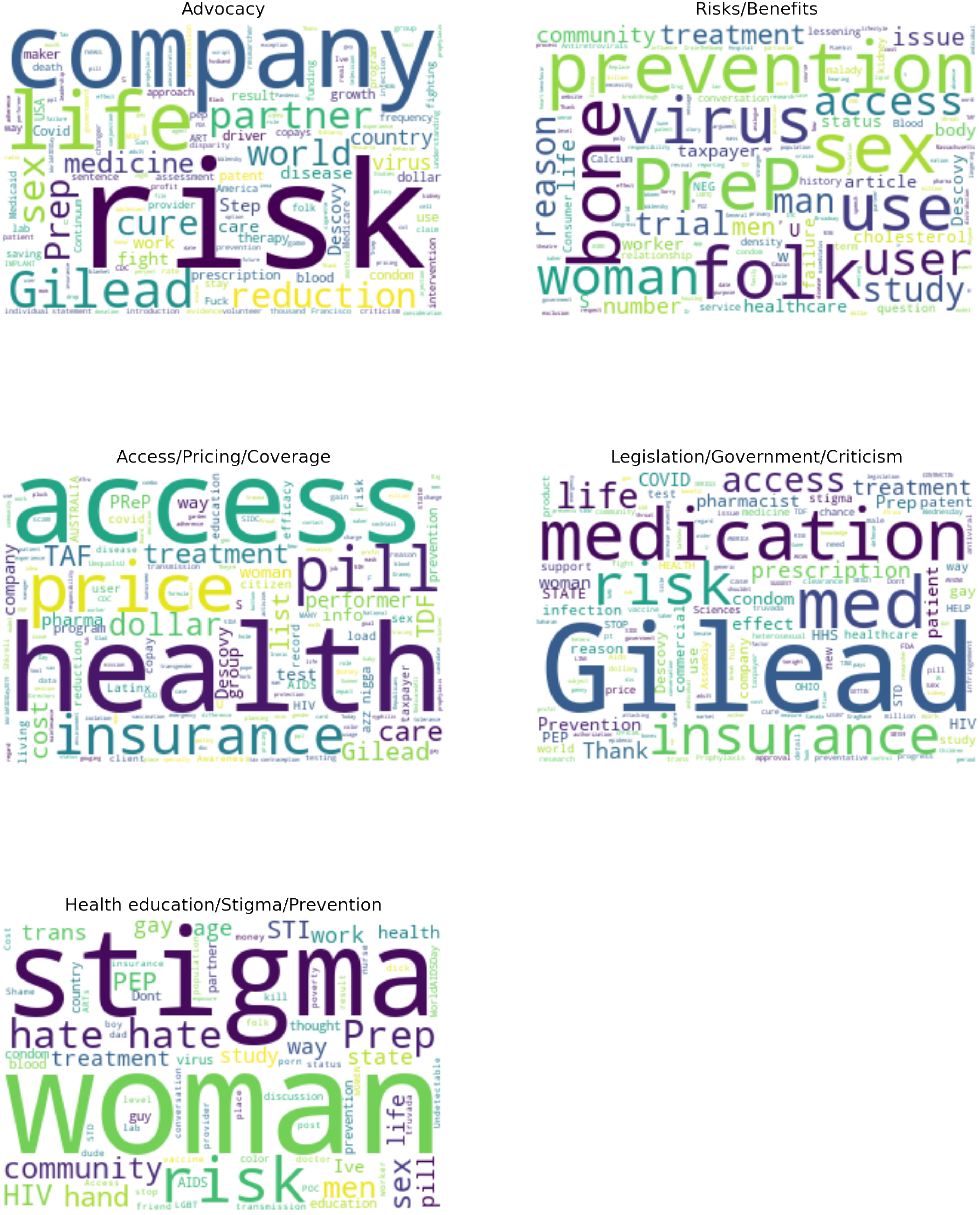
Word clouds of the topics in topic modeling

Topic 1 is the advocacy about medicines, drugs, drug companies (e.g., Gilead), health care.

> *Example tweet 1: “The American people hold a patent on an HIV prevention drug that could save thousands of lives, but instead the government is letting a private company profit from it. It’s time for the CDC to assert its patent and make PrEP affordable for all*.*”*

> *Example tweet 2: “Maybe the USA should advertise about Truvada instead of beer and cigarettes but the USA is the only country in the world that allows prescription drugs to be advertised on television radio so we pay much more than any other country does for medicine is that confident leadership?”*

Topic 2 is about risks and benefits involving Truvada, drugs, PrEP, etc.

> *Example tweet 1: “Also for my folks taking Truvada for prevention or treatment, there have been studies suggesting the lessening of bone density with long term use. So vitamin D and Calcium are super important!”*

> *Example tweet 2: “I have a few lingering questions*… *Is Truvada safe enough for OTC distribution? Is the new formulation with the tenofovir analogue (Descovy)*

> *significantly better than Truvada w*.*r*.*t. side effects, or is this mainly for patent purposes? And who funded the clinical trials?”*

Topic 3 is about the access, pricing, and insurance coverage of Truvada/PrEP.

> *Example tweet 1: “Meanwhile Truvada for Prep is $60*.*00 per pill (approximately); a drug that was researched and developed with taxpayer dollars. Health insurance companies and citizens are being price gouged by big pharma on a drug that cost them next to nothing to develop*.*”*

> *Example tweet 2: “It’s hivpreventionday so it’s an important time to remind everyone that PrEp, one of the biggest breakthroughs in HIV prevention costs $1300 a month when the pills cost dollars to make. The gov. neglected the AIDS crisis before and capitalizing on it now isn’t out of character*.*”*

Topic 4 is about legislation, criticism, complains about the HIV, PrEP.

> *Example tweet 1: “The Assembly just passed my bill (SB1021) to cap prescription drug co-pays; ensure people can actually use their insurance to obtain life-saving medication. The bill also ensures that insurance compa-nies donate undermine access to PrEP (a powerful HIV preventative). Thank you!”*

> *Example tweet 2: “Less than 1 in 4 Americans who could benefit from Pre-Exposure Prophylaxis are on it, with communities of color, women people living in the South underrepresented among PrEP users. Tell Congress to make PrEP affordable available to all! “*

Topic 5 mainly talks about the health education, prevention and stigma about HIV and PrEP.

> *Example tweet 1: “I don’t understand this tweet tbh. After all the medical evidence and science behind U=U the efficacy of PrEP and us trying to lift the stigma related to HIV in our community what is your fear about working with undetectable performers? “*

> *Example tweet 2: “Apparently with the exception of myself every friend and mu closest friends*.. *I know are engaging in raw sex with random dudes*… *Learn to love yourselves and place value for your life. Geez. As a former Hiv counselor*… *The virus stills infects and kills. Get on prep if negative*.*”*

## IV. Discussion and Conclusions

With the advent of Web 2.0, social media has been widely used for health-related purposes such as health interventions, medical education, and disease outbreak surveillance [20]. More importantly, patients, especially those who are young, are often connected with social media. It is estimated that 80% of cancer patients use social media to connect with peers [21]. User-generated social media posts such as tweets can provide insights about the public’s perception, cognitive, and behavioral responses to health-related issues [20]. At the same time, they can be used to disseminate health information and combat misinformation. PrEP, as one of the most effective ways to reduce the risk of HIV infection, has been widely discussed on social media. Despite the effectiveness of PrEP in the prevention of HIV transmission, its utilization is low in the US [22]. The lack of uptake is especially low among populations disproportionately affected by HIV such as the age group of under 24 years old [23]. It is therefore important to understand the barriers to the wider use of PrEP in the US. In this study, to identify such barriers, we focused on tweets about personal discussions. With machine learning, we were able to use PubMedBERT-abstract-only model to classify the tweets into three categories: personal discussion, promotional information, and irrelevant with micro F1 score of 0.842. Among the personal discussion tweets, the issues related to PrEP/Truvada include advocacy, risks/benefits, access, pricing, insurance coverage, legislation, stigma, health education, and prevention of HIV. This finding is consistent with a recent narrative review about the barriers to the wide use of PrEP (i.e., knowledge/awareness of PrEP, perception of HIV risk, stigma from healthcare providers or family/partners/friends, distrust of healthcare providers/systems, access to PrEP, costs of PrEP, and concerns around PrEP side effects/medication interactions) [22]. In addition, it appears that pricing is the major concern that impedes the wide use of PrEP. Many people advocate for better access to PrEP with insurance coverage. Therefore, public and private insurers should reduce or eliminate copay to increase access. For people without health insurance, they should be directed to medication assistance programs of both governments at different levels and pharmaceutical manufacturers (e.g., https://www.cdc.gov/hiv/basics/prep/paying-for-prep/insured.html).

## Data Availability

The study used openly available data from Twitter.

## A. Limitations

A few limitations need to be noted in this study. First, we only included the tweets between 2018 and 2022. The reason is that in May 2018, FDA approved expanded indication for Truvada for reducing the risk of acquiring HIV-1 in adolescents [24]. Second, we used only the MeSH terminology for term analysis. This could have resulted in some missed terms/synonyms or misspelling used in tweets.

## B.Future work

In future work, we will use the results of both topic modeling and content analysis to develop strategies for promoting PrEP to young adults who are at risk or with HIV. We will also use geo-tagged tweets to identify risk behaviors in tweets such as alcohol intake and assess their impact on the uptake of PrEP and HIV endemic.

## V.Acknowledgements

We would like to thank undergraduate students Jessica Valyou and Sarah Helgeson for their efforts in manually labelling the tweets. We would also like to thank undergraduate students Neissa Philemon and Jessica Valyou for their help with the literature review of this project.

## Footnotes

This study was partially supported by the National Institute on Alcohol and Alcoholism under award number P01AA029547, and University of Florida-Florida State University Clinical and Translational Science Award, which is supported in part by the National Center for Advancing Translational Sciences under award number UL1TR001427.

## Notes

### Competing Interest Statement

The authors have declared no competing interest.

### Author Declarations

The study used openly available data from Twitter.

## References

[1] E. Walsh-Buhi, R. F. Houghton, C. Lange, R. Hockensmith, J. Ferrand, L. Martinez, et al., “Pre-exposure prophylaxis (prep) information on instagram: Content analysis,” JMIR Public Health and Surveillance, vol. 7, no. 7, e23876, 2021.

[2] L. A. Stutts, P. A. Robinson, B. Witt, and D. F. Terrell, “Lost in translation: College students’ knowledge of hiv and prep in relation to their sexual health behaviors,” Journal of American College Health, vol. 70, no. 2, pp. 561–567, 2022.

[3] S. Z. Kudrati, K. Hayashi, and T. Taggart, “Social media & prep: A systematic review of social media campaigns to increase prep awareness & uptake among young black and latinx msm and women,” AIDS and Behavior, vol. 25, no. 12, pp. 4225–4234, 2021.

[4] P. S. Loosier, K. Renfro, M. Carry, S. P. Williams, M. Hogben, and S. Aral, “Reddit on prep: Posts about pre-exposure prophylaxis for hiv from reddit users, 2014– 2019,” AIDS and Behavior, vol. 26, no. 4, pp. 1084–1094, 2022.

[5] J. J. Matacotta, F. J. Rosales-Perez, and C. M. Carrillo, “Hiv preexposure prophylaxis and treatment as prevention—beliefs and access barriers in men who have sex with men (msm) and transgender women: A systematic review,” Journal of Patient-Centered Research and Reviews, vol. 7, no. 3, p. 265, 2020.

[6] I. Gregory Phillips, A. B. Raman, D. Felt, et al., “Prep4love: The role of messaging and prevention advocacy in prep attitudes, perceptions, and uptake among ymsm and transgender women,” Journal of acquired immune deficiency syndromes (1999), vol. 83, no. 5, p. 450, 2020.

[7] X. Yang, T. Fang, S. A. Mobarak, et al., “Social network strategy as a promising intervention to better reach key populations for promoting hiv prevention: A systematic review and meta-analysis,” Sexually Transmitted Infections, vol. 96, no. 7, pp. 485–491, 2020.

[8] A.-M. D. Navarra, C. Handschuh, T. Hroncich, S. K. Jacobs, and L. Goldsamt, “Recruitment of us adolescents and young adults (aya) into human immunodeficiency virus (hiv)–related behavioral research studies: A scoping review,” Current HIV/AIDS Reports, vol. 17, no. 6, pp. 615–631, 2020.

[9] A. van Heerden and S. Young, “Use of social media big data as a novel hiv surveillance tool in south africa,” Plos one, vol. 15, no. 10, e0239304, 2020.

[10] S. D. Young, C. Rivers, and B. Lewis, “Methods of using real-time social media technologies for detection and remote monitoring of hiv outcomes,” Preventive medicine, vol. 63, pp. 112–115, 2014.

[11] C. Liu and X. Lu, “Analyzing hidden populations online: Topic, emotion, and social network of hiv-related users in the largest chinese online community,” BMC medical informatics and decision making, vol. 18, no. 1, pp. 1–10, 2018.

[12] M. L. McHugh, “Interrater reliability: The kappa statistic,” Biochemia medica, vol. 22, no. 3, pp. 276–282, 2012.

[13] J. Devlin, M.-W. Chang, K. Lee, and K. Toutanova, “Bert: Pre-training of deep bidirectional transformers for language understanding,” arXiv preprint 1810.04805, 2018.

[14] J. Lee, W. Yoon, S. Kim, et al., “Biobert: A pretrained biomedical language representation model for biomedical text mining,” Bioinformatics, vol. 36, no. 4, pp. 1234–1240, 2020.

[15] Y. Peng, S. Yan, and Z. Lu, “Transfer learning in biomedical natural language processing: An evaluation of bert and elmo on ten benchmarking datasets,” arXiv preprint 1906.05474, 2019.

[16] Y. Gu, R. Tinn, H. Cheng, et al., Domain-specific language model pretraining for biomedical natural language processing, 2020. xeprint: 2007.15779.

[17] L. Soldaini and N. Goharian, “Quickumls: A fast, unsupervised approach for medical concept extraction,” in MedIR workshop, sigir, 2016, pp. 1–4.

[18] D. M. Blei, A. Y. Ng, and M. I. Jordan, “Latent dirichlet allocation,” Journal of machine Learning research, vol. 3, no. Jan, pp. 993–1022, 2003.

[19] R. Rehurek and P. Sojka, “Gensim–python framework for vector space modelling,” NLP Centre, Faculty of Informatics, Masaryk University, Brno, Czech Republic, vol. 3, no. 2, 2011.

[20] J. Chen, Y. Wang, et al., “Social media use for health purposes: Systematic review,” Journal of medical Internet research, vol. 23, no. 5, e17917, 2021.

[21] L. A. Braun, B. Zomorodbakhsch, C. Keinki, and J. Huebner, “Information needs, communication and usage of social media by cancer patients and their relatives,” Journal of Cancer Research and Clinical Oncology, vol. 145, no. 7, pp. 1865–1875, 2019.

[22] K. H. Mayer, A. Agwu, and D. Malebranche, “Barriers to the wider use of pre-exposure prophylaxis in the united states: A narrative review,” Advances in Therapy, vol. 37, no. 5, pp. 1778–1811, 2020.

[23] P. S. Sullivan, R. M. Giler, F. Mouhanna, et al., “Trends in the use of oral emtricitabine/tenofovir disoproxil fumarate for pre-exposure prophylaxis against hiv infection, united states, 2012–2017,” Annals of epidemiology, vol. 28, no. 12, pp. 833–840, 2018.

[24] Gilead, U.S. Food and Drug Administration Approves Expanded Indication for Truvada® (Emtricitabine and Tenofovir Disoproxil Fumarate) for Reducing the Risk of Acquiring HIV-1 in Adolescents, https://www.gilead.com/news-and-press/press-room/press-releases/2018/5/us-food-and-drug-administration-approves-expanded-indication-for-truvada-emtricitabine-and-tenofovir-disoproxil-fumarate-for-reducing-the-risk-of-/, [Online; accessed 18-Aug-2022], 2018.

